# An interpretable cell-centric representation of pancreatic ductal adenocarcinoma matches multiple-instance learning for clinical outcomes and leads to novel pathologist-driven histological biomarkers

**DOI:** 10.64898/2026.10.03.26364596

**Authors:** Maxence Pelloux, Catherine TL Tran, Jumanah Baig, Abdelhakim Khellaf, Sally Lang Huang, Olivier Tung Nguyen, Maëlle Batardière, Sarah-Slim Diwan, Zhiyuan Yang, Jia Lin Li, Vyky Vu, Simon F Roy, Geneviève Soucy, Bich Ngoc Nguyen, Basile Tessier-Cloutier, Mahdi S Hosseini, David JHF Knapp, Séverine Valmary-Degano, Vincent QH Trinh

**Affiliations:** Centre Hospitalier Universitaire Grenoble Alpes, Grenoble, France; Centre Hospitalier de l’Université de Montréal, Montreal, Quebec, Canada; Institute for Research in Immunology and Cancer, Université de Montréal, Montreal, Quebec, Canada; Research Institute of the McGill University Health Center, Montreal, Quebec, Canada; Gina Cody School of Engineering and Computer Science, Concordia University, Montreal, Quebec, Canada; Division of Dermatology and Department of Pathology, McGill University Health Centre, Montreal, Quebec, Canada; Rosalind and Morris Goodman Cancer Institute, McGill University, Montreal, Quebec, Canada; Département de pathologie et biologie cellulaire, Université de Montréal, Montreal, Quebec, Canada; University Grenoble Alpes, Inserm UMR 1209 / CNRS 5309, Institute for Advanced Biosciences, Department of Pathology, Grenoble Alpes University Hospital, Grenoble, France; Centre de recherche du Centre Hospitalier de l’Université de Montréal, Montreal, Quebec, Canada

## Abstract

Current state of the art computational pathology foundation models attain satisfying accuracy most general pathology tasks including detection of pancreatic ductal adenocarcinoma (PDAC) but fail to provide sufficient explainability, operating as “black boxes”. Some interpretability effort has been made, with models providing attention maps that project back on the slide the models high attended areas that pushed for the given prediction result, without further naming or reasoning explanation which can limit pathologists trust toward the prediction. We developed C3PRO, a cell-centric framework attempting to provide deeper understanding and explainability, by using embeddings extracted using a pathology foundation model, from cell centered tiles containing both the cell and its close neighborhood, which were then clustered into recurrent morphological phenotypes. A spatial graph was additionally constructed on top of the back-projected vocabulary to account for recurrent motifs. The evaluation of the model performance involved a cohort of 809 patients (825 diagnostic biopsies) using nested cross validation. C3PRO was benchmarked against four different multiple instance learning frameworks and achieved on-par performance for diagnostic (0.958 AUC), survival (0.617) and resectability (0.696) prediction, while providing additional insights compared to the strongest baseline models. Each clustered phenotype was probed and extracted as a representative tile mosaic and annotated by two expert pathologists, constituting a phenotype vocabulary available for statistical analysis. These phenotypes were scored by pathologists with physical brightfield microscopy and replicated the independent statistical significance impact on overall survival prediction from diagnostic biopsies.

**STATEMENT OF SIGNIFICANCE:** Pathology foundation model multiple-instance-learning frameworks cannot be audited by the pathologists. We demonstrate that a representation constructed from identifiable cell phenotypes is on par with state-of-the-art models for diagnosis, prognosis, and resectability in pancreatic cancer. This approach also uncovers prognostic stromal states that expert observers cannot differentiate using light microscopy.

## INTRODUCTION

Pancreatic ductal adenocarcinoma (PDAC) is infamously known as part of the most lethal solid malignancies, with a five year overall survival under 13% across all stages,^1^ mostly explained by late symptomatic manifestation, aggressive clinical behavior and resistance to systemic therapy.^2^ As for most malignant entities pathology plays a key three-step role the care of these patients: diagnosis on the often limited biopsy material, prognostic stratification (although very limited) to guide the intensity and timing of therapy, and participation in the assessment of resectability. Each of these steps reported to being subject to substantial inter-observer variability, whether in grading biopsies, assessment of precursor lesions or tumor regression grading on resection specimen after neoadjuvant treatment.^3^ Some attempts to improve or provide additional input to prognostic prediction was made through deep-learning models built on conventional clinico-pathological parameters, but these rarely exceed a concordance index of 0.70.^4^ Objective, reproducible and interpretable computational tools are needed.

Digital whole-slide imaging (WSI) has enabled the application of deep learning to pathology, at first via fully-supervised convolutional neural networks (CNN) that required substantial and costly expert annotation,^5^ then through weakly supervised frameworks such as multiple-instance learning (MIL) operating at the slide level with no need for extensive pixel-level annotations.^6^ These pipelines rely on pathology foundation models, pre trained on large datasets using self-supervision, such as UNI,^7^ Virchow,^8^ Prov-GigaPath^9^ and H-Optimus-0,^10^ which are transferable and now used to produce embeddings for a wide range of downstream tasks such as slide-level diagnostic and prognostic predictions.

In order to provide these slide level predictions, the information carried by the extracted tile-embeddings representing a fraction of the overall tissue must be aggregated using different approaches such as attention-based MIL,^11^ transformer architectures,^12^ or state-space models.^13^ Graph neural networks (GNN), that build graph dependencies on cellular or tile level representation encode tissue architecture and cellular connections explicitly,^14^ which present as a good fit for cell-centered representations given that the composition and structure of the tumor microenvironment shape PDAC behavior.^15^

Independently of the aggregation method, these models fail to provide clear explainability and predictions are difficult for pathologists to verify. A first solution serving as the main interpretive tool is the attention heatmap, which comes with limitations, as attention is an imperfect proxy for prediction drivers,^16^ and do not or partially account for biological reasoning. Fundamentally, a heatmap signals without naming: showing high yield prediction areas without elaborating on their content. As an example a high attention region for the diagnosis of most malignancy could include neoplastic epithelium, desmoplastic stroma or an immune infiltrate, which all carry different implication. Furthermore, a model whose reasoning cannot be probed is difficult to audit for systematic errors or to integrate into practice especially in a field such as pathology where the responsibility relies on the practitioner and not the model, therefore requiring in order for the model to be adopted in daily routine practice.

Cell centered approaches also offer a path to improved interpretability, by changing the unit of analysis from an arbitrary grid applied to the tissue slide to tiles centered on a biological nameable entity.^17^ Clustered into recurrent phenotypes these presentations can then be inspected, and therefore annotated, constructing a vocabulary in the spirit of concept-based models,^18^ that emerge unsupervised from the data. These morphological phenotypes could also for example relate to molecular subtypes of PDAC, which carry prognostic value^19^ and are known to have detectable morphological signatures on hematoxylin-eosin sections.^20^

We introduce C3PRO (Computational Cell-Centric Prognosis and Resectability Outcome) a novel cell-centric approach that builds an interpretable representation of the tissue from clustered foundation-model embeddings extracted from cell-level tiles centered on detected nuclei centroids, reflecting the given cell and its close neighborhood. The pipeline then builds a vocabulary of recurrent cell phenotypes using clustering methods followed by a spatial graph constructed on top of the back-propagated clusters (Figure 1). We make three contributions: Applied to PDAC biopsy digital WSIs, (1) C3PRO matches state-of-the-art MIL approaches for diagnosis, prognosis and resectability, providing higher interpretability while preserving equal performance. (2) Each of the clustered cell-phenotype is displayed as a mosaic of representative cells and annotated by two expert-pathologists, resulting in a morphological vocabulary which is then confronted with conventional histopathological review of the digital slides. (3) Downstream statistical analysis identifies cellular phenotypes associated with survival and resectability. It also shows that recurrent spatial neighborhoods provide no additional prognostic information beyond cellular composition.

**Figure 1.**
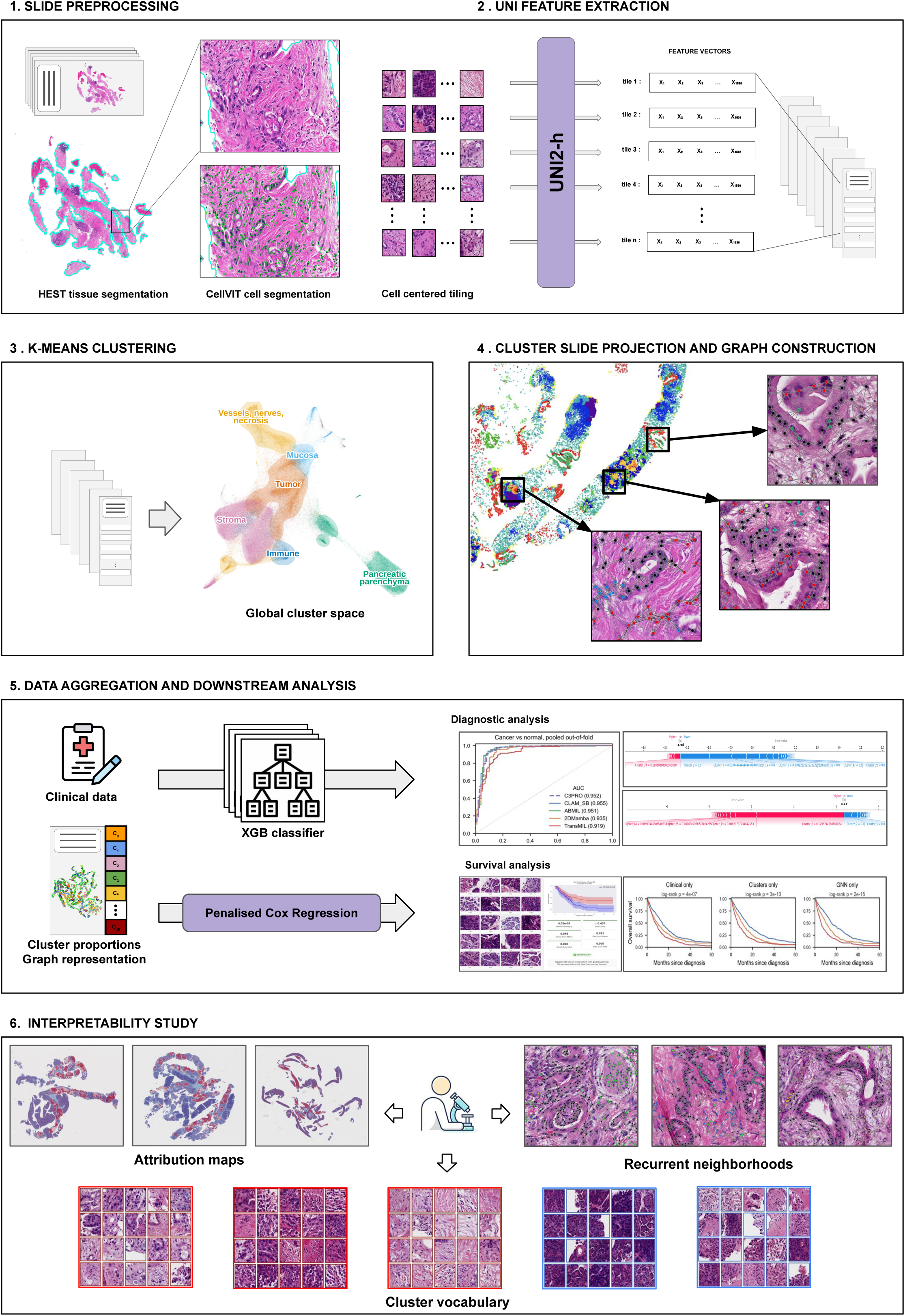
Overview of the C3PRO Pipeline. 1. **Tissue Segmentation and Nucleus Detection:** Tissue is segmented on the whole-slide image, and individual nuclei are detected and segmented. 2. **Encoding Cells:** A tile centered on each nucleus centroid is encoded by a pathology foundation model into a 1536-dimensional vector, representing each cell with its immediate tissue context. 3. **Phenotype Clustering:** These embedding vectors are partitioned using spherical K-means into a vocabulary of fifty recurrent cell phenotypes. The map, an illustration, is a UMAP of 247,463 cells (300 per slide) coloured by tissue compartment; shaded areas contain 85% of the cells of each compartment. 4. **Phenotype Assignment and Graph Construction:** Each cell is assigned its phenotype and projected back onto the slide, and a spatial graph links each cell to its nearest neighbors. 5. **Downstream analyses:** Slides are represented by their phenotype proportions and a graph embedding, combined with clinical covariates, and passed to penalized models under patient-grouped nested cross-validation: gradient boosting for binary tasks and penalized Cox regression for survival. 6. **Prediction Inspection:** Since every feature is a cell phenotype, predictions can be inspected. Attributions are projected back onto the slide, each phenotype is displayed as a mosaic of its constituent cells for pathological annotation, and recurrent spatial neighborhoods are examined.

## MATERIALS AND METHODS

### Study cohort

We assembled a retrospective cohort of pancreatic biopsy whole-slide images, all from the Centre Hospitalier de l’Université de Montréal (CHUM), from a registry of 1791 patients who were subject to pancreatic core needle biopsy. We applied the following exclusion criteria: absence of essential clinical data, non-PDAC histological type (including neuroendocrine tumors and other non-ductal neoplasms), pre-invasive neoplastic lesions without carcinoma (including IPMN and PanIN), and non-representative samples with insufficient tissue for histological analysis. The resulting cohort consisted of 809 patients with 825 slides. Hematoxylin-eosin-stained biopsies were sampled by endoscopic ultrasound-guided techniques (EUS-FNB) and scanned at 40x magnification.

One important detail is that the slides covered two different acquisition eras corresponding to a change of slide-scanner between 2021 and 2022, which will be referred subsequently as “pre-scanner” era: comprising 635 patient biopsies acquired from 2017 to 2021; and “post-scanner” era: with 174 patients ranging from 2022 to 2024. Therefore, a systematic source of appearance variation was anticipated and accounted for in both fold construction and clustering, as detailed below. All clinical, pathological and survival follow-up data were obtained under protocol 2025-12351, approved by the Centre de recherche du Centre Hospitalier de l’Université de Montréal. Cohort characteristics are summarized in Table 1.

**TABLE 1.** Characteristics of the study cohort (n = 809)

| **Characteristic** | **Patients, n (%)** |
|---|---|
| Slides | 825 (16 patients with two slides) |
| **Acquisition period** |  |
| 2017–2021 (first scanner) | 635 (78.5) |
| 2022–2024 (second scanner) | 174 (21.5) |
| Age at diagnosis, years, median (IQR) | 70.3 (63.4–76.7) |
| Missing | 80 (9.9) |
| Male sex | 449 (55.5) |
| **Histological grade on biopsy** |  |
| G1 | 162 (20.0) |
| G2 | 431 (53.3) |
| G3 | 181 (22.4) |
| Registry code 0 | 8 (1.0) |
| Missing | 27 (3.3) |
| **Tumor location** |  |
| Head | 503 (62.2) |
| Body | 204 (25.2) |
| Tail | 70 (8.7) |
| Diffuse (head, body and tail) | 3 (0.4) |
| Missing | 29 (3.6) |
| **Large-vessel involvement on pre-treatment imaging** |  |
| Present | 336 (41.5) |
| Absent | 440 (54.4) |
| Other registry code | 1 (0.1) |
| Missing | 32 (4.0) |
| Tumor size on pre-treatment imaging, cm, median (IQR) | 3.0 (2.5–3.8) |
| Missing or non-numeric | 50 (6.2) |
| Surgical resection | 171 (21.1) |
| Missing | 1 (0.1) |
| Neoadjuvant FOLFIRINOX* | 125 (15.5) |
| Deaths | 726 (89.7) |
| Follow-up, months, median (reverse Kaplan–Meier) | 55.8 |
| Overall survival, months, median (95% CI) (Kaplan–Meier) | 10.3 (9.3–11.4) |

Percentages are given over all 809 patients. \*Recorded in the registry only when administered. IQR, interquartile range; CI, confidence interval.

### Cell detection and tile-embedding

For each whole slide image, the cellular nuclei were segmented using CellViT++,^17^ a foundation-model-based segmentation pipeline based on a Segment Anything Model backbone. The slides were processed at 40× magnification with an overlapped 1,024 × 1,024-pixel tile grid (including a 128-pixel tile overlap) resulting in per-slide nucleus centroids and contours as the output. For each detected cell a 224 × 224-pixel vignette centered on the nucleus centroid was extracted from the slide and embedded using UNI2-h,^7^ a pathology foundation model pre-trained on over 100,000 whole-slide images. Each tile was thus converted into a 1,536-dimension embedding vector capturing a given cell morphology and its local microenvironment. Because adjacent cells share part of their vignette, their embeddings are expected to covary. Across the cohort, this generated approximately 41 million cellular embeddings.

### Cell phenotype vocabulary

The different cellular phenotypes were built by clustering the single-cell feature embeddings using spherical K-means (with K = 50, chosen as described below, and 15 epochs). Two separate clustering spaces were built for each of the two differing cohorts (the prognosis cohort consisting of invasive carcinoma only, and the diagnosis sub-cohort comprising additional slides who were diagnosed as negative for PDAC). This ensured that the cluster indices were specific to each cohort and not comparable across them. Each vocabulary was fitted on the slides of the task it serves, so that its fifty clusters are spent on the morphological variability of that population. Spherical K-Means was implemented using L2-normalized feature vectors, preventing cluster assignment to be influenced by embedding magnitude, and making it sensitive only to direction in the 1536D spherical space. Thus, removing the inductive bias induced by foundation-model representation whose norm tend to be influenced by the pre-training objective. The number of desired phenotypes was determined a priori and not tuned on the endpoints, ensuring sufficient phenotypic granularity while taking into account the need for each cluster to be subsequently reviewable by expert pathologists. A post hoc sweep analysis was still performed with K values ranging from 10 to 100, on the diagnostic vocabulary, and using criteria blind to all clinical endpoints. It found no optimum value with silhouette and clustering stability both decreasing linearly with K, and silhouette remaining close to zero throughout the sweep (0.047 to 0.070), which is an expected behavior for such a continuous embedding space. Therefore, the chosen K value reflects a granularity decision rather than the recovery of a discrete number of cell types as you would expect with transcriptomics. K = 50 fell within the stability plateau and produced the most robust granularity across clusters. The same value was applied to the prognostic vocabulary, for which the K sweep was not performed due to insufficient computational resources. Domain shift was also considered to prevent appearance shifts between scanner eras, by applying mean-centering per era before normalization. The streaming K-means method is mathematically equivalent to batch K-means, and processes one slide at a time so the full corpus of 41 million cells for the prognostic cohort, and 12 million cells for the diagnosis subcohort, never completely resides in memory. The output of this step is, for each slide, a 50-dimension vector of cluster proportions which is used as bag-level features for downstream classification and prediction tasks, along with per-cell cluster assignments to build the spatial graphs.

### Spatial graph and survival model

A spatial cell graph was built for each slide using PyTorch, with each detected cell nucleus serving as a graph node, with a one-hot encoding of its phenotype cluster. The graph edges connected each cellular unit to its six nearest neighbors on the digital slide image, thus capturing the local cellular neighborhood. A graph attention network (GAT) was trained on top of these graphs to predict a per-slide hazard score using a Cox partial-likelihood target. The network was made up of three attention convolutional layers: two attention heads in the first two layers and one in the third; with ELU activation and dropout; followed by global mean pooling to convert the output into a 32-dimensional slide-level embedding; which was finally passed through a two-layer perceptron to produce a log-hazard. The model was optimized using Adam with a cosine-annealing schedule, with early stopping on a held-out validation subset. The 32-dimensional slide embeddings generated by the attention model were then used as image-derived features for the downstream survival analyses.

### Diagnostic model

For the diagnostic task, which involved separating cancer from non-cancer tissue, a post-scanner sub-cohort of 294 slides (195 malignant and 99 benign, from 274 patients) was used to build a specific phenotype vocabulary. Gradient boosting was applied to the slide-level cluster proportions vectors to predict the diagnosis, which allowed us to model per-cluster contributions with SHAP values. Cross-validation was performed with patient-grouping to ensure that slides from the same patient did not appear in both training and evaluation folds (some patients having multiple slides tied to them).

### Multiple instance learning baselines

Comparison of C3PRO’s performance with other weakly supervised frameworks was performed against four different well established architectural baselines: an attention-based MIL (ABMIL),^11^ a clustering constrained attention MIL (CLAM),^21^ a transformer based model (TransMIL),^12^ and a state space model (2DMamba).^13^ For each of these baselines the same foundation model was used (UNI2-h), and evaluation was carried on the same patient folds. The main difference between the four baselines and our cell-centric pipeline is the construction of the tiles which correspond to an arbitrary grid that covers the entire tissue. Using the same foundation model and data partition means that performance variation can most likely be attributed to the representation of the data and its aggregation rather than encoder, cohort or data split differences. For each baseline a per-slide risk score was computed for survival and evaluated against the cell-centric models using the same concordance-index convention.

### Evaluation and statistical analysis

For prognosis and resectability, 5x5 cross-validation was used with the same 25 patient folds applied to every model. To avoid patient leakage, we made sure that all slides from a given patient would end up in the same fold by generating folds grouped by patient identifier. We also made sure to balance the two scanner eras by stratifying by acquisition year. For diagnosis prediction we used a separate partition (StratifiedGroupKFold, grouped by patient identifier). Two design features were specifically introduced to account for optimistic bias: (1) the GNN output vectors were generated out-of-fold, meaning that for each of the twenty-five folds a separate network was trained on the folds specific training set, guarantying that no test patient would contribute to the GNN output used to evaluate it. All feature standardization and model fitting were similarly confined to the training split of each fold.

The metrics used for performance evaluation were Harrell’s concordance index averaged across the twenty-five folds and generated per fold on the held-out test set; Area under the ROC curve for diagnostic and resectability prediction. Model comparisons were carried out using Wilcoxon test on paired scores with Holm correction applied for multiple comparisons. Patients were split into risk tertiles based on prediction scores. Survival was compared using the log-rank test.

### Recurrent motif discovery

Each phenotypic entity (cluster) was depicted as a mosaic composed of tile samples representative of all slides including the cluster with a minimum radius separation between selected samples. This ensured both inter- and intra-slide representativity, making sure that the tile mosaic would span across the whole cohort rather than a few slides. Once sampled, the fifty mosaics were reviewed by independent expert pathologists blinded to survival curves and effect, characterizing the dominant cellular type and the architectural pattern.

We also explored whether recurrent spatial motifs carried additional survival insights, in a similar manner as the cellular-neighborhood framework introduced by multiplexed imaging.^22^ We retrieved for each cell the ten nearest neighbors and their cluster were extracted and summarized in a proportion vector of the fifty phenotypes. These neighborhoods were clustered across all cells and all slides using K-means into ten recurrent neighborhoods which were then used to build neighborhood composition vectors for each slide. and tested against overall survival using Cox regression with and without adjustment for the six clinical variables and Benjamini-Hochberg correction applied.

Because a neighborhood dominated by one cellular phenotype would carry redundant information as that phenotype’s abundance, every significant neighborhood was tested with a Cox model with the slide proportion of its dominant cellular cluster, to distinguish genuine effect carried by architecture versus redundant effect from cellular composition. The neighborhood size was also varied through six values (four, six, ten, twenty, thirty, and fifty nearest cells) while keeping a fixed number of neighborhoods. Benjamini-Hochberg correction was also applied across all tests.

### Conventional histopathological review

To confront our vocabulary to routine practice clinical criteria an expert pathologist reviewed 466 slides and evaluated five criteria (examples in Supplementary Figure S1): preserved acinar parenchyma, tumor cellularity, stromal morphology, lymphocytic infiltrate and tumor necrosis. We matched vocabulary entities to their corresponding criteria and fixed them before any analysis of the readers annotations: per-slide summed proportions of the acinar, neoplastic and immune phenotypes, and for stromal morphologies the share of dense stroma in a maximum 400-pixel radius around tumor cells. Tumor necrosis was added after the initial analysis of the other four criteria; its counterpart is the only phenotype whose descriptions by both readers mention necrosis. Concordance was computed with Spearman correlation for relative values and area under the ROC curve for ordinal and binary criteria. Each criterion was evaluated alongside the six clinical variables for prognosis, with nested Cox models on the survival folds to compare with the phenotype proportions.

### Implementation

The pipeline was implemented in Python, using CellViT++ for cell detection, UNI2-h for feature extraction, a GPU streaming spherical K-means for clustering, PyTorch Geometric for the graph attention network, and scikit-survival, scikit-learn, XGBoost and SHAP for modeling and interpretation. All stochastic components used a fixed seed.

## RESULTS

### Cohort characteristics

The cohort consisted of 809 patients with pancreatic ductal adenocarcinoma (consisting of 825 slides), from a unique center. The median age at diagnosis was 70.3 years (IQR 63.4–76.7), with a slight male predominance at 55.5%. Most tumors were moderately differentiated (grade 2, 53.3%), 20.0% were grade 1 and 22.4% were grade 3. The majority of tumors were located in the pancreatic head (62.2%), 25.2% originated from the body and 8.7% from the tail. 41.5% of cases reported large vessel involvement, in line with the fact that only 21.1% were subsequently subject to surgical resection. The median tumor size was 3.0 cm (IQR 2.5–3.8). Neoadjuvant therapy (FOLFIRINOX) was administered to 15.5% of patients. Median follow-up was 55.8 months, 89.7% of patients had died, and the median overall survival was 10.3 months (95% CI 9.3–11.4). This coincides with the known high rate of mortality, mostly imputable to the late-stage, unresectable nature of the PDAC population. Some clinical and biological variables carrying known prognostic value in PDAC such as CA 19-9, were not uniformly recorded and are thus not reported; while some variables were not included by choice, due to their posteriori nature in order to fairly reproduce the setting of the pathologist reading the biopsy sample (pathological TNM stage, nodal status, resection and margin status).

### Diagnostic performance and its morphological basis

C3PRO achieved a per fold mean area under the ROC curve of 0.958 (standard deviation 0.043; pooled out-of-fold 0.952) for the diagnostic task, with a sensitivity of 0.964 and specificity of 0.838 at a probability threshold of 0.5. The MIL baselines reached a similar performance range (pooled out-of-fold: CLAM 0.955, ABMIL 0.951, 2DMamba 0.935, TransMIL 0.919), with sensitivities and specificities at the same threshold of 0.954 and 0.869 (CLAM), 0.949 and 0.869 (ABMIL), 0.959 and 0.818 (2DMamba), and 0.985 and 0.576 (TransMIL). Since the diagnostic partition was split in five folds the per-fold paired testing is inadequate and underpowered; hence the pooled out-of-fold discrimination was directly compared using the DeLong test on the 294 slides. Our cell centered approach was statistically indistinguishable from 3 baselines: CLAM (Figure 2A; ΔAUC −0.002), ABMIL (ΔAUC 0.002), and 2DMamba (ΔAUC 0.017), staying non-significant after Holm correction; and significantly surpassed only TransMIL. The cell-centric representation therefore matched the strongest MIL baselines without penalty, while making its decision inspectable, as detailed below.

**Figure 2.**
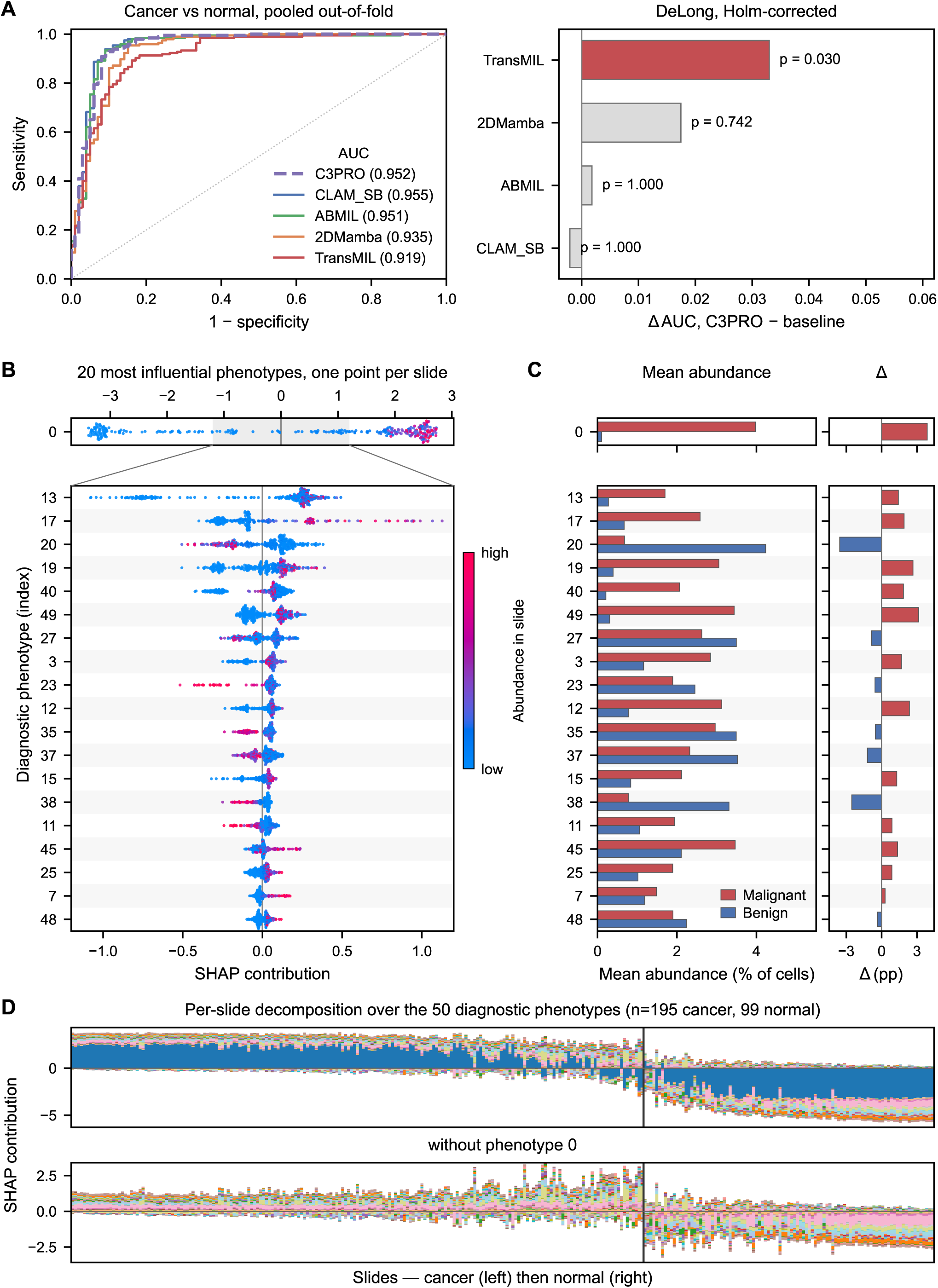
Diagnostic Performance and Its Morphological Basis. **(A)** ROC curves for malignant versus benign slides, from pooled out-of-fold predictions on the 294-slide sub-cohort (left), and differences in area under the curve between C3PRO and each multiple-instance learning baseline, trained on the same patient-grouped folds with the same foundation model, with Holm-corrected DeLong p values (right). C3PRO was statistically indistinguishable from CLAM, ABMIL and 2DMamba and exceeded TransMIL (p = 0.030). **(B)** SHAP attribution of the twenty phenotypes with the highest mean absolute SHAP value, one point per slide, colored by the abundance of the phenotype in the slide (blue, low; red, high). The leading phenotype (top strip, full scale) spans roughly −3.4 to +2.7, whereas the other nineteen (below, axis enlarged to the shaded ±1.2 interval) all stay within ±1.13: the prediction rests largely on a single phenotype. **(C)** Mean abundance of the same twenty phenotypes (percentage of cells) in malignant and benign slides, and their difference; rows are aligned with panel B. The leading phenotype rises from 0.1% of cells in benign tissue to 4.0% in malignant tissue, while the fourth moves in the opposite direction. **(D)** Per-slide decomposition of the diagnostic prediction into the signed contributions of the fifty phenotypes, slides ordered from malignant to benign; the leading phenotype is the only one drawn in dark blue. The lower band shows the same contributions without the leading phenotype, on its own scale (model not refitted). Phenotypes are identified by index, as the diagnostic vocabulary is fitted separately from the prognostic one and is not annotated.

Additionally, the cluster-based nature of the pipeline makes the diagnostic prediction inspectable, first by using the SHAP scores of the proportion model, which allows the identification of strong separators of malignancy. The diagnostic task was dominated by a single phenotype contributing six times more than the next cluster with a secondary tier of phenotypes carrying the remaining discriminative power (Figure 2B-D; Figure 3). Taking advantage of this, we observe that malignancy drivers are composed of carcinomatous structures, on desmoplastic stroma, and on necrotic areas, reproducing key diagnostic cues used in routine practice, while preserved pancreatic parenchyma, both exocrine and endocrine, mature stroma, and other incidental structures such as vessels and duodenal mucosa contribute in the opposite direction. Another reassuring observation is that uncommon histological types such as signet-ring cell carcinoma and undifferentiated carcinoma, also were strong predictors of malignancy even though they were less represented in the dataset, meaning that prediction is not confined to the conventional ductal morphology. Lastly, we also notice that chronic pancreatitis is not attributed to malignancy prediction, another reassuring fact since it is the principal differential diagnosis and a classical source of false positives on biopsy material. Refitting the diagnostic model without the most discriminating phenotype still maintained high performance with a slightly lowered area under the curve from 0.958 to 0.938 per fold (0.952 to 0.923; DeLong p = 0.017), the remaining phenotypes still carried most of the discrimination (Figure 2D).

**Figure 3.**
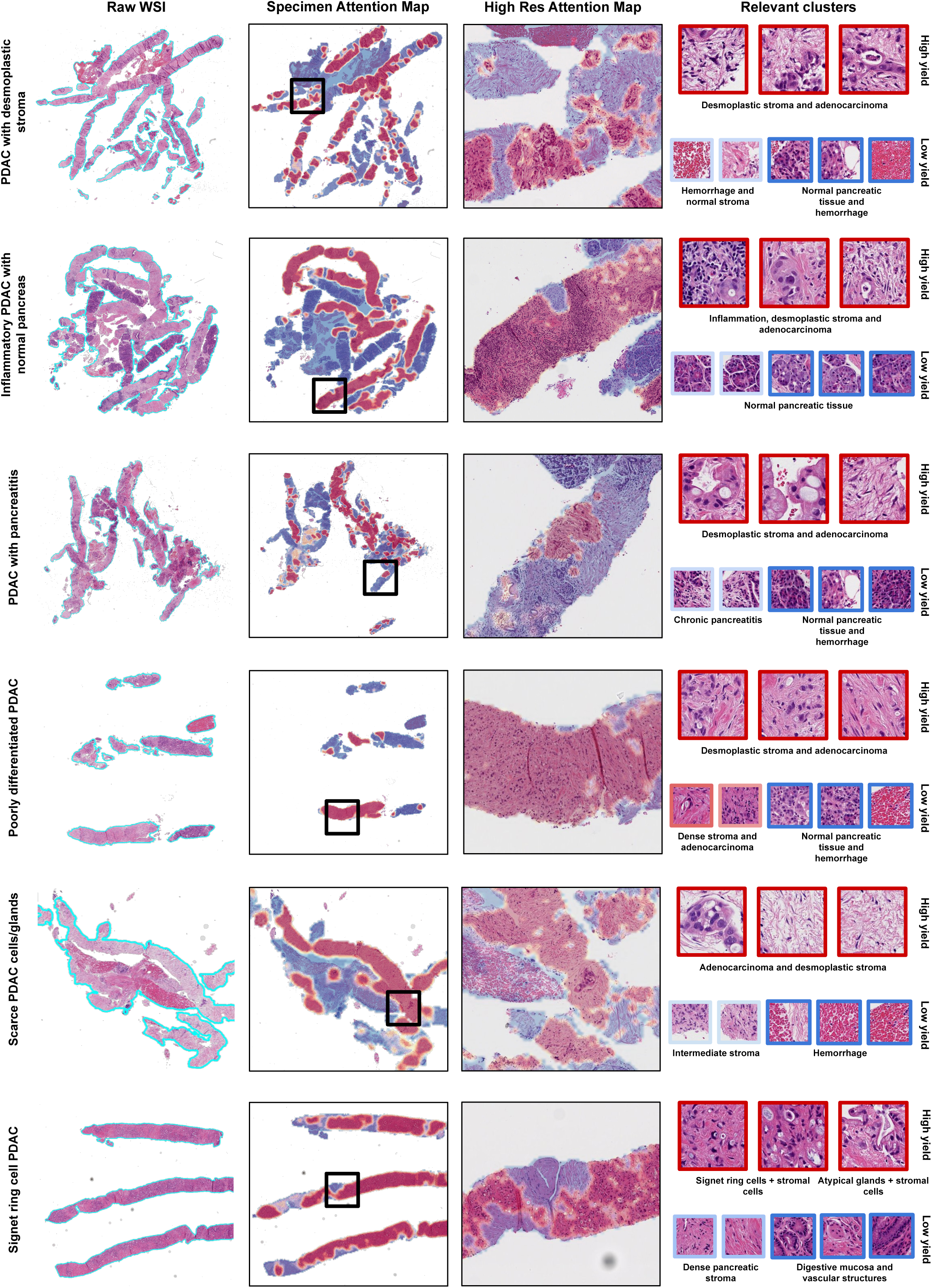
Case-Level Interpretability of the Diagnostic Model. Six biopsy cases illustrating the range of morphologies encountered in routine practice: ductal adenocarcinoma with desmoplastic stroma, inflammatory adenocarcinoma with residual normal pancreas, adenocarcinoma arising on chronic pancreatitis, poorly differentiated adenocarcinoma, a specimen with scarce neoplastic glands, and signet-ring cell adenocarcinoma. From left to right: hematoxylin-eosin slide; attribution map at specimen scale; high magnification of the boxed region; phenotypes contributing most to the prediction, each shown by a sample with its pathological description, outlined in red when contributing towards malignancy and in blue when contributing in the opposite direction. Attribution concentrates on neoplastic glands and their desmoplastic stroma in all six cases, including the poorly differentiated and signet-ring variants, and is directed away from residual normal parenchyma, haemorrhage and chronic pancreatitis.

### Prognostic performance: parity with MIL

The overall survival was modeled in 809 patients with 5×5 nested cross-validation. The clinical-only model used six clinical variables only available at biopsy, excluding surgical status which is unknown at diagnosis and mediates prognosis rather than predicting it. This clinical baseline achieved a concordance index of 0.575. Adding the graph embedding raised it to 0.617, an increment of 0.042 present in 24 of the 25 folds (Figure 4A). Both image-only representations exceeded the clinical baseline (cluster proportions 0.588, graph embedding 0.602; Figure 4B). Adding the surgical status improved the best model concordance to 0.671, as previously said, a post-biopsy variable and only used as a sensitivity analysis.

**Figure 4.**
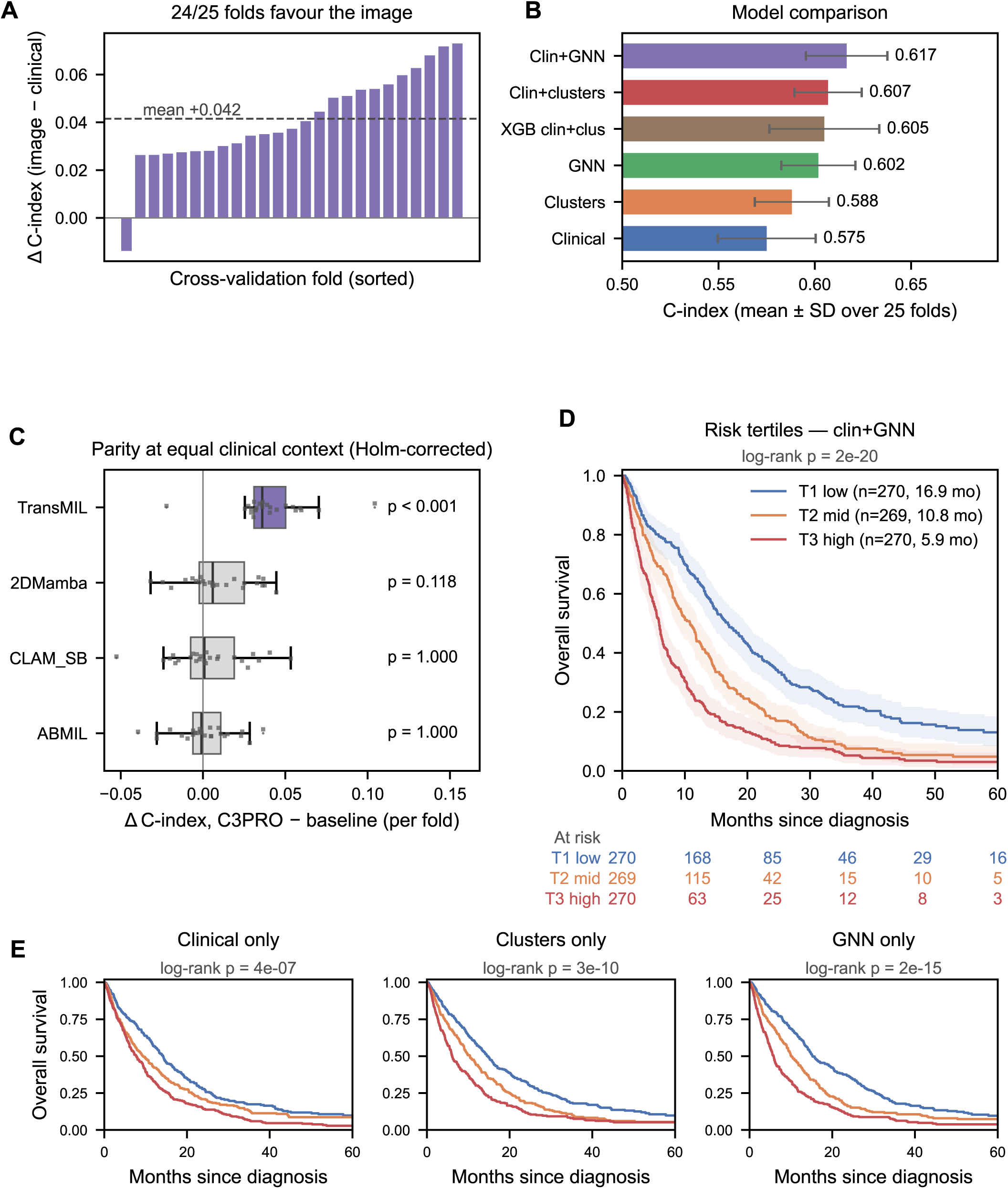
Prognostic Performance of the Cell-Centric Representation. **(A)** Difference in concordance index between the champion model (six biopsy-available clinical variables and the graph embedding) and the clinical variables alone, in each of the 25 folds of the 5×5 nested cross-validation; positive in 24 folds (mean +0.042, dashed line). **(B)** Concordance index of the six prognostic models (mean ± standard deviation over the 25 folds): 0.575 for the clinical variables available at biopsy, higher for both image-only representations, and 0.617 for the clinical variables combined with the graph embedding. **(C)** Per-fold difference in concordance index relative to the champion, computed per patient, for each multiple-instance learning baseline combined with the same six clinical variables on identical folds, individual folds overlaid. Differences from ABMIL, CLAM and 2DMamba were not significant after Holm correction; only TransMIL differed (Holm-adjusted p < 0.001). **(D)** Kaplan–Meier overall survival by tertile of the champion model’s out-of-fold risk score, with numbers at risk. Median survival was 16.9, 10.8 and 5.9 months from the lowest to the highest risk tertile (global log-rank p = 2×10^-20^; all pairwise comparisons significant). **(E)** The same stratification applied to each feature block alone: all three blocks separated the tertiles, with an amplitude following the concordance index of the underlying model.

Against MIL baselines, with concordance computed per patient under the same conditions, the image-only graph embedding (0.607) was comparable to 3 of the 4 baselines except TransMIL, which it exceeded (ABMIL 0.604, CLAM 0.603, 2DMamba 0.595, TransMIL 0.540); cluster proportions alone (0.593) were slightly below CLAM (−0.010, p = 0.041) and ABMIL (−0.011, p = 0.059). Adding the six clinical variables did not change the observed pattern (C3PRO 0.622, ABMIL 0.621, CLAM 0.618, 2DMamba 0.612, TransMIL 0.581; Figure 4C) with C3PRO matching state-of-the-art MIL, while representing a framework more open to pathological inspection.

Combining the clinical variables with the graph embedding outperformed their combination with the cluster proportions by 0.010 (0.617 versus 0.607; 18 of 25 folds), while adding the graph on top of the proportions added only 0.002 (19 of 25 folds): the information carried by the graph overlaps substantially with the proportions. This is consistent with the quasi-null result obtained in the recurrent neighborhoods search. The proportional hazards assumption was not rejected for the champion model (global Schoenfeld p = 0.80; tumor size and large vessel involvement deviated individually); a gradient-boosting model using the six clinical variables and cluster proportions, free of that assumption, reached a similar concordance (0.605 against 0.607 for the Cox model on the same inputs).

### Prognostic stratification

Patients were stratified into risk groups based on the out of fold risk score tertile acquired from the champion model, which created clear separate clinical risk groups with median overall survival of 16.9, 10.8, and 5.9 months (Figure 4D; global log-rank p = 2×10^-20^). Statistical significance was reached for all three pair comparisons, which for example included the high risk versus intermediate risk comparison, therefore, showing clear separation between the groups rather than an artificial sub-stratification of a uniform poor prognosis group. Even though this stratification is retrospectively determined, it still points that the model can identify prognostic subgroups with clinical relevance.

This three-strata separation was also performed on each component of the champion model (e.g. clinical variables alone, cluster proportions alone, and graph embeddings alone); and found that each separated the cohort in distinct tertiles (Figure 4E), with the two non-clinical components producing marginally wider separations. The combined clinical-plus-graph model produced the widest separation of all.

### Resectability

With a prevalence of 20.7%, resectability prediction was evaluated on the same cohort and folds as the prognostic prediction. The clinical-variables-alone model reached an AUC of 0.681, and adding the graph embeddings improved it to 0.696. The image-only models were weaker (cluster proportions 0.644, graph embedding 0.627) and the four MIL baselines weaker still (ABMIL 0.613, CLAM 0.614, 2DMamba 0.578, TransMIL 0.551). Again, C3PRO was at parity with the strongest baselines and beat only TransMIL. The graph embeddings and cluster proportions performed near-identically (0.696 versus 0.697). Again, the morphological added value remained small, which is expected for a decision that mostly relies on radiological criteria.

### Interpretability: phenotype clusters and recurrent motifs

All fifty morphological phenotypes were displayed as mosaics, composed of representative cluster tile samples, and subsequently characterized by two expert pathologists blinded to the survival and resectability associations. Without specifically specifying annotation categories, we observed a high level of agreement in the terminology used by both reviewers for 11 of the significant 14 clusters: such as neoplastic epithelium, desmoplastic stroma, and normal exocrine parenchyma. Where the labels differed, the focus was on the level of detail rather than the crude identity of the phenotype: one reviewer differentiated desmoplastic from collagenous fibrosis, or immature from mature stroma, where the other used a single term.

Fourteen clusters were significantly associated with overall survival after adjustment for the clinical covariates (Figure 5), with a stable effect for all clusters. With surgical status added as a seventh covariate, seven phenotypes were significant, six of them among the fourteen. The adverse prognosis clusters included multiple sub-phenotypes of carcinoma and stroma: Poorly differentiated (hazard ratio 1.20), discohesive tumor cells (1.10) and moderately differentiated carcinoma (1.20), were associated with worse prognosis. This predictive contribution is additional to the histological grade which was already included in the clinical covariates. Well differentiated PDAC also pushed toward adverse outcome but did not reach statistical significance (1.07, q = 0.19). On the opposite end of the spectrum, the protective phenotypes included preserved exocrine and acinar parenchyma (0.88 and 0.89). In contrast, a chronic pancreatitis phenotype rich in immune cells was adverse (hazard ratio 1.11, 95% CI 1.03–1.20), and remained so after adjustment for tumor burden, the summed proportion of the tumor phenotypes (1.11, 1.04–1.20). An emphasis should be made about the stromal phenotypes, since entities were identified on both ends of the predictive spectrum: some adverse (1.14, 1.12, 1.10) and some protective phenotypes (0.87, 0.87, 0.89). While one pathologist gave most of the stromal phenotypes the same label, the second characterized two of the pejoratives as desmoplastic and called the protective ones loose, mixed and immune-rich stroma, hinting at the poorly documented and reproducible prognosis influence of the stromal compartment maturity.

**Figure 5.**
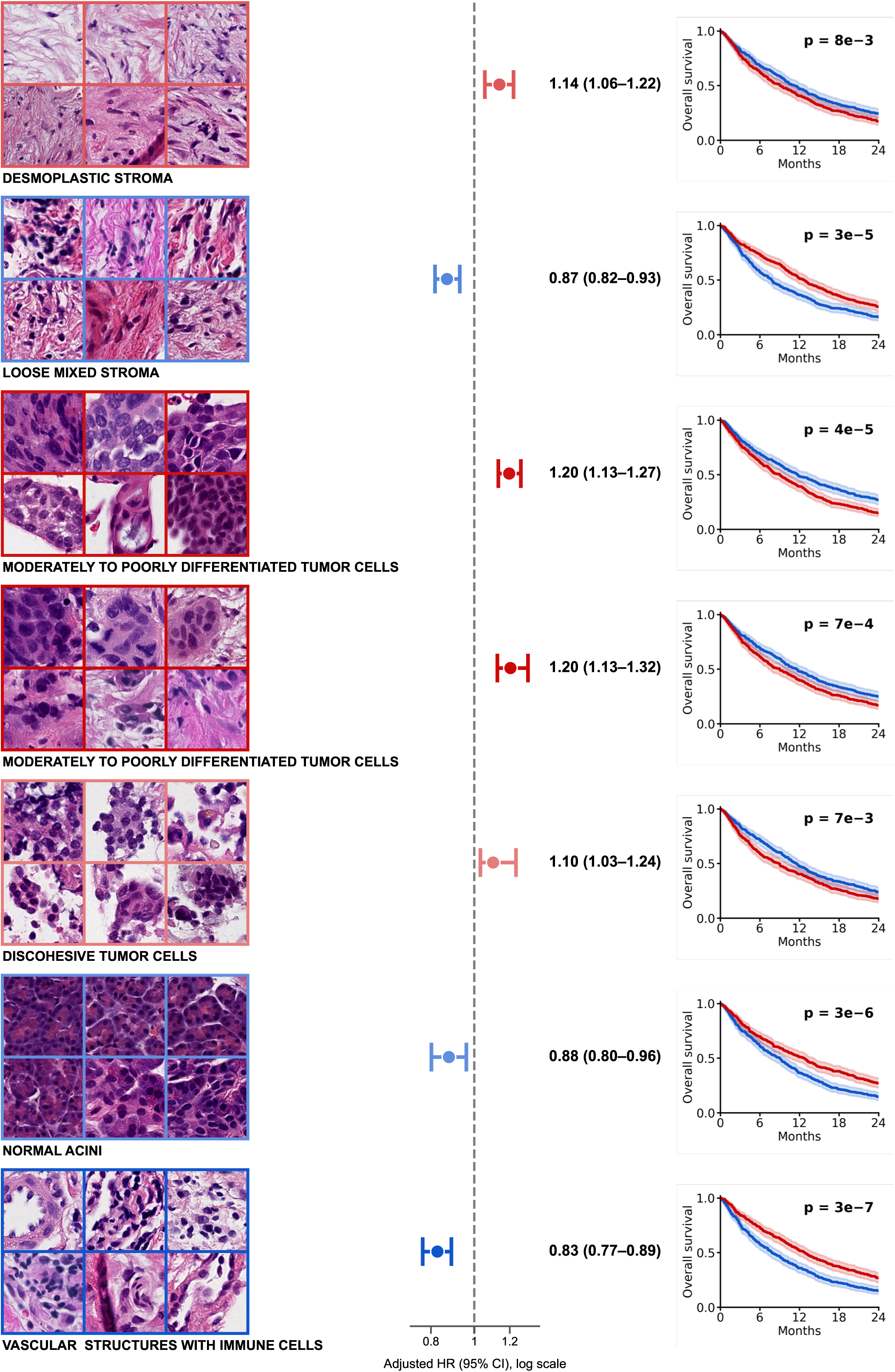
Morphological Identity of the Prognostic Phenotypes. Seven of the fourteen phenotypes significantly associated with overall survival, chosen to cover their morphological diversity. Rows 1–2, two stromal phenotypes of opposite prognostic direction (desmoplastic stroma, adverse; loose mixed stroma, protective). Rows 3–5, carcinoma (two phenotypes of moderately to poorly differentiated tumor cells, and discohesive tumor cells). Row 6, normal acini. Row 7, vascular structures with immune cells. **Left Panel:** Six representative cells per phenotype, sampled across the cohort, framed by color according to the adjusted hazard ratio (red, adverse; blue, protective). **Center Panel:** Adjusted hazard ratio for a one-standard-deviation increase in the phenotype’s slide proportion, with 95% confidence interval, on a logarithmic axis. **Right Panel:** Kaplan–Meier overall survival for slides above (red) and below (blue) the cohort median proportion of the phenotype, with 95% confidence bands, truncated at 24 months, and log-rank p value. These curves are univariate and descriptive, whereas the hazard ratios are adjusted, so the two can diverge. Each carcinoma phenotype shown was adverse, and the first reader described the two stromal phenotypes of rows 1 and 2 in nearly identical terms despite their opposite effects. The last phenotype was also associated with resection (adjusted odds ratio 1.32, q = 0.021).

Regarding resectability, two clusters were in line with their prognostic value and showed significant effect after clinical adjustment, supporting the biological rather than incidental nature of these associations: The first cluster consisted of poorly differentiated carcinoma and was associated with lower odds of resection (0.61 OR) and shorter survival (1.20); the second significant cluster, vascular structures with immune cells, was associated with both increased odds of resection (1.32) and longer survival (0.83).

The study of recurrent spatial neighborhoods in contrast found almost no additive prognostic value compared to the crude phenotypic composition. Thirty-one neighborhoods were significantly associated with survival across the six neighborhood-sizes and two different vocabulary granularities (K = 10 and K = 50 phenotypes) that we tested. 30 of the 31 neighborhoods were highly dominated by a unique cluster and were statistically indistinguishable from the proportion of that given cluster regarding prognosis. The remaining neighborhood carried a modest prognostic association (1.10 HR). Therefore indicating that most of the predictive signal coming from this cohort resided in the tissue composition rather than the spatial relation between the different phenotypes.

### Conventional histopathological criteria against the phenotype vocabulary

We asked one expert pathologist to review and characterize quantifiable and qualitative routine histopathological criteria for 441 patients of our main cohort, showing no detectable difference in survival (median of 10.3 months in both datasets, log-rank p = 0.81) but a higher share of low grade carcinoma (26% against 13% of grade 1 PDAC; p < 0.001). We observed high agreement on pathologist-model paired quantitative morphological counterparts, ρ = 0.89 for areas characterized as preserved acinar parenchyma, and 0.68 for tumor cellularity, and moderate agreement for ordinal or binary criteria, with areas under the curve of 0.80 for necrosis, 0.71 for dense stroma, 0.67 for immature stroma, and 0.67 for lymphocyte density (κ 0.35, 0.34 and 0.31 for necrosis, stroma and lymphocytes; Figure 6).

**Figure 6.**
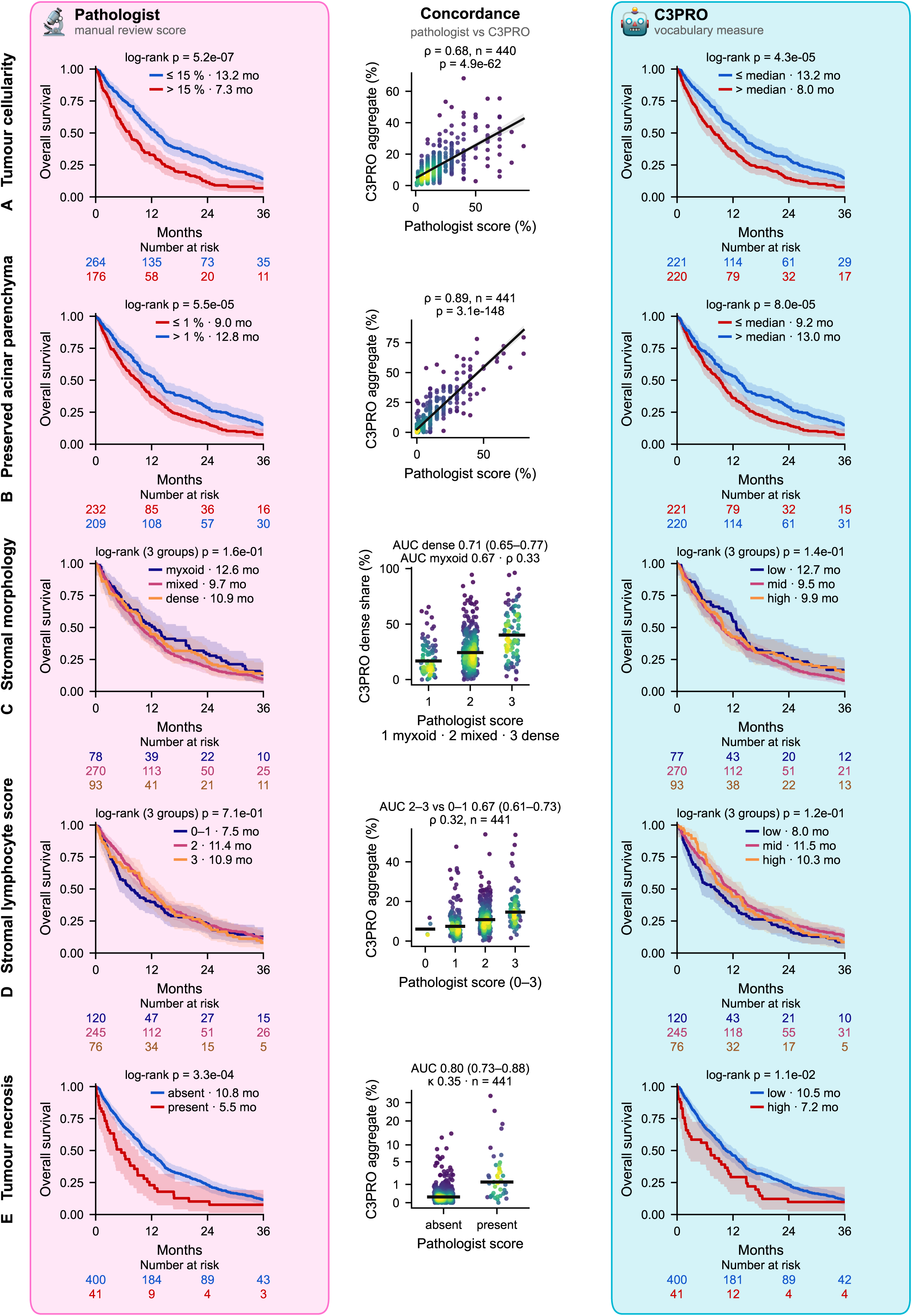
Conventional histopathological review against the C3PRO phenotype vocabulary. Five criteria scored by one pathologist on one slide per patient are compared with C3PRO measures. Each row shows overall survival by the pathologist’s score (left), the concordance between the two (centre) and overall survival by the C3PRO measure (right). **(A)** Tumour cellularity. **(B)** Preserved acinar parenchyma. **(C)** Stromal morphology, from 1 (myxoid) to 3 (dense collagenous). **(D)** Lymphocytic infiltrate adjacent to tumour cells, from 0 to 3. **(E)** Tumour necrosis. Survival: Kaplan–Meier estimates with 95% confidence bands and log-rank p values; A and B are split at the median, E by presence, and C and D by score level (0 and 1 pooled in D), with C3PRO groups of the same sizes; red, shorter survival (A, B, E). Concordance: Spearman’s ρ (A, B); area under the ROC curve with 95% confidence interval for dense stroma, lymphocyte score 2– 3 and necrosis, followed by ρ (C, D) or Cohen’s κ (E). Square-root axis in E; point colour shows local density.

After clinical adjustment tumor cellularity (hazard ratio 1.31, 95% CI 1.19–1.45) and preserved acinar parenchyma (0.86, 0.78–0.96) remained significant for prognosis, lymphocyte density did not. Necrosis had the strongest effect (hazard ratio 1.91, 1.37–2.66; median survival 5.5 against 10.8 months) which also persisted after adjustment for tumor cellularity. The stromal compartment showed heterogeneous prediction, with mixed stroma being the most adverse pattern, without significant prognostic group separation (log-rank p = 0.16). The same observation was made for the matched vocabulary groups. In nested Cox models, the five criteria raised the concordance index of the clinical model from 0.582 to 0.624, and the phenotype proportions to 0.603. On this subset, five expert-scored criteria captured at least as much prognostic information as the phenotype proportions. The graph embedding, however, combined with the same clinical variables, matched the expert criteria (0.628) and added a small increment beyond them (0.010; 18 of 25 folds; Holm p = 0.019).

## DISCUSSION

In this study, we aimed to explore whether a cell-centric representation of tissue slides could match traditional and state of the art multiple instance learning pipelines across diagnosis, prognosis and resectability prediction, using the same UNI2-h encoder, while providing improved reasoning transparency for the pathologist. Our framework achieved performance statistically indistinguishable from three MIL baselines and exceeded the fourth. We did not aim to surpass them but instead aimed toward increased routine practice value through legibility by producing decomposable and auditable predictions. Since the same UNI2-h encoder was used, the key distinguishing factor for this representation is explainability, our cell-centric method provides a morphological vocabulary alongside prediction score for each entity it comprises. This is a substantial shift compared to the baselines whose explainability relies mostly on attention heatmaps. In diagnosis, the prediction was dominated by a single phenotype and a small secondary tier. This is also consistent with the biological reasoning that pathologists use in routine practice, with diagnostic decision focusing on a rather small number of histological diagnostic criteria, it also illustrates the high-interpretability nature of our method, being allowed to decompose the model reasoning rather than only displaying an opaque attention map back onto the slide (Figure 2B-D; Figure 3). The prediction did not, however, depend on that phenotype alone: without it, performance remained high, the remaining phenotypes carrying most of the discrimination.

This architecture also allowed us to explore further into the spatial cellular arrangement field through the use of graph neural networks and found limited prognostic additive value beyond crude phenotypic composition. The graph embedding added 0.042 of C-index to clinical variables but only a marginal 0.010 to cluster composition, and constituted our champion model for prognosis; for resectability it matched the cluster proportions (0.696 against 0.697). Of the 31 recurrent niches that were significant for prognosis, across six neighborhood scales, 30 were redundant with the proportion of the dominant phenotype. This should be interpreted as specific to our setting (i.e. PDAC biopsies from a single center) and not as a general law; and within it, cellular composition was the main carrier of prognostic information rather than spatial arrangement of the tissue. Each vignette, although centered on a cell, already captures much of its immediate surroundings, and the six nearest neighbors often lie within overlapping vignettes, so that the graph largely re-encoded a local field already present in the embeddings. Larger-scale architecture was not explored, the largest neighborhood tested comprising fifty cells, and may still carry prognostic information, which would require dedicated features such as tissue topology. On the subset of patients reviewed by a pathologist, the graph embedding matched five expert-scored criteria, each combined with the clinical variables (0.628 against 0.624), and added a small increment beyond them (0.010; 18 of 25 folds), which phenotype proportions did not.

Fourteen phenotypes were associated with prognosis after adjustment for clinical covariates, deliberately excluding surgical status in order to maintain an identical setting to that of practicing pathologists receiving a biopsy (seven phenotypes were significant when it was added); resectability associations were almost entirely explained by clinical confounding. They closely reproduced established pathology criteria with neoplastic epithelium and desmoplastic stroma carrying adverse prognosis weight, and preserved acinar parenchyma being protective. All the above associations are in line with basic pathology reasoning, where replacement of functional parenchyma by tumor and desmoplasia is synonym of disease progression. To our surprise, one phenotype corresponding to chronic pancreatitis, carried an adverse prognosis (HR 1.11), independently of tumor burden, which could perhaps reflect obstructive pancreatitis upstream of the tumor.

Beyond these expected associations, two takeaways add to the established pathology criteria. First, the neoplastic epithelium remained adverse even though histological grade was included in the adjusted covariates. This was the case for moderately and poorly differentiated PDAC, as well as discohesive tumor cells. The well-differentiated phenotype however had a smaller, non-significant effect, consistent with the known prognostic value of the differentiation gradient. Our representation adds the quantity of tumor in the sampled tissue as a prognostic factor, which conventional reporting does not record. This is consistent with the conventional review, in which the pathologist-scored tumor cellularity remained prognostic after clinical adjustment (hazard ratio 1.31). Second, the stromal compartment contained phenotypes showing opposite prognostic direction, that the first reader described almost identically, while the second called two of the adverse ones desmoplastic and the protective ones loose, mixed or immune-rich. This is reminiscent of the recognized but poorly reproducible grading of stromal maturity.^23,24^ With these observations we can generate the hypothesis that the embedding space captures stromal states that are not entirely recognized by routine pathology description, pointing towards the need for deeper molecular stromal subtyping and validation. This was also supported by the conventional histopathology review, where the association of stromal morphology with survival was heterogeneous, with mixed stroma (which was implemented as the “intermediate” stromal category) being the most adverse pattern, and remained non-significant on the reviewed subset. Thus, a three-tier grading for stromal morphology fits poorly with the survival pattern observed here, supporting the hypothesis that further immunohistochemical or molecular validation is needed for the stromal compartment.

Against the conventional review, the cluster vocabulary closely reproduced the pathologist’s quantitative scores (acinar parenchyma and tumor cellularity), and moderately the ordinal ones, without annotation. It carried no more prognostic value than the five criteria evaluated by the expert pathologist, which bounds the overall vocabulary contribution. The vocabulary does not discover additional prognostic information; it offers the expert’s reading at the scale of a whole cohort, with finer resolution.

These results are subject to several limitations, first the cohort was built from a single institution, external validation on independent cohorts is needed to show whether our method is robust to scanner and center variations. In particular, the per-era mean-centering would have to be re-estimated on the slides of any new scanner or center. Second, the conventional histopathological review was carried out by a single pathologist, aware of the study, which was also involved in the analysis of the phenotypes annotation, therefore these results should mostly be interpreted for agreement evaluation rather than independent validation. Third the clinical model lacks several variables of established prognostic value such as CA 19-9 levels, formal TNM stage and nodal status, that were not uniformly available, therefore undermining the clinical variable’s prognosis performance. Fourth, the paired tests were performed on overlapping cross-validation folds, which means that the 25 measured C-index are not independent, although leaving the parity conclusions intact. Fifth, the feature embeddings used to build the vocabulary were extracted and annotated on this same cohort, even though unsupervised and never using the outcomes, it should be replicated on independent data. Sixth, embedding each of the 41 million cells is computationally heavier than tile-level multiple-instance learning, which adds computing time, energy use and environmental cost, to be weighed against the expected benefit, particularly for diagnosis, where conventional reading is already fast and accurate.

Future perspective and on-going work include external validation which constitutes the immediate priority, followed by molecular profiling of the phenotypes and correlation with already known molecular subclasses of PDAC such as basal-like and classical molecular subtypes, through spatial transcriptomics.

Finally, no component of the framework is tied to the pancreas, each processing step can be applied to any tissue, and a new vocabulary will be learned without prior labeling. The downstream prediction layers are also standard models. Therefore, the same approach could be applied to any organ, disease or clinical question.

## CONCLUSION

A representation of pancreatic histology built from individual cells, their foundation-model embeddings and a vocabulary of recurrent phenotypes predicts diagnosis, survival and resectability as well as attention-, transformer- and state-space-based multiple-instance learning, while every feature it uses is a phenotype a pathologist can view and name. Parity is its precondition, interpretability its contribution. What it reveals is modest in effect size, coherent with established pathology, and in the stroma more finely resolved than routine description. External validation is required before any clinical claim, but a model whose reasoning can be expressed in the vocabulary of morphology is one whose errors a pathologist can audit and challenge.

## Data Availability

Please contact the corresponding author for data access. WSIs can be shared through material transfer agreements between research institutions. The code of the C3PRO pipeline is available at https://github.com/PathSpace/c3pro_pancreas.; the scripts generating the figures and tables are available from the corresponding author on request.

## ACKNOWLEDGMENTS

Please contact the corresponding author for data access. WSIs can be shared through material transfer agreements between research institutions. The code of the C3PRO pipeline is available at https://github.com/PathSpace/c3pro_pancreas; the scripts generating the figures and tables are available from the corresponding author on request. VQT is funded by the Fonds de recherche Quebec – Santé, the Canadian Cancer Society, the Université de Montréal, the National Science and Engineering Council of Canada, MedTeq+, the Institute for Research in Immunology and Cancer, the Fonds de recherche Quebec – Nature et technologies, and the Canadian Foundation for Innovation. SFR holds a clinical research scholar award from the Fonds de recherche Quebec – Santé. MB was supported by the Fonds Pierre-Saul. Claude (Anthropic) was used to assist with code development and editing of parts of the manuscript text. All outputs were reviewed and verified by the authors, who take full responsibility for the content.

---

**Supplementary Figure S1.**
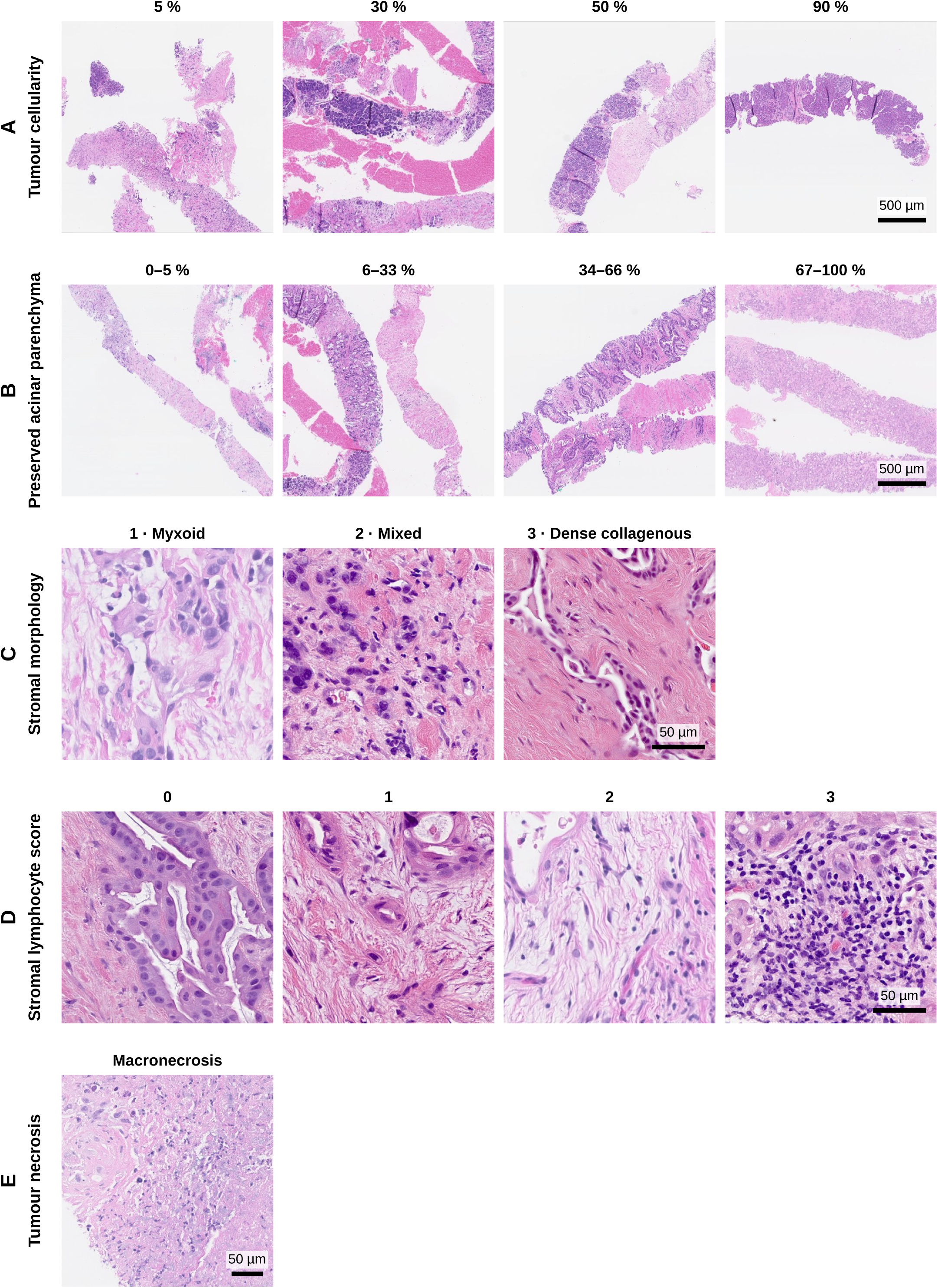
Criteria of the conventional histopathological review. Representative examples, hematoxylin and eosin. **(A)** Tumour cellularity, at 5, 30, 50 and 90%. **(B)** Preserved acinar parenchyma, from 0–5% to 67–100%. **(C)** Stromal morphology: predominantly myxoid (1), mixed (2) and predominantly dense collagenous (3). **(D)** Lymphocytic infiltrate adjacent to tumour cells, scores 0 to 3. **(E)** Macronecrosis. Within each row, all images cover the same field of view (A and B, 2.2 mm; C and D, 200 µm; E, 340 µm); scale bars, 500 µm in A and B and 50 µm in C to E.

## REFERENCES

1. Siegel RL, Giaquinto AN, Jemal A. Cancer statistics, 2024. CA Cancer J Clin. 2024;74(1):12-49.

2. Mizrahi JD, Surana R, Valle JW, Shroff RT. Pancreatic cancer. Lancet. 2020;395(10242):2008–2020.

3. N Kalimuthu S, Serra S, Dhani N, et al. Regression grading in neoadjuvant treated pancreatic cancer: an interobserver study. J Clin Pathol. 2017;70(3):237–243. doi:10.1136/jclinpath-2016-203947

4. Xu D, Zhang K, Li M, et al. Prognostic nomogram for resected pancreatic adenocarcinoma: a TRIPOD-compliant retrospective long-term survival analysis. World J Surg. 2020;44(4):1260–1269.

5. Coudray N, Ocampo PS, Sakellaropoulos T, et al. Classification and mutation prediction from non-small cell lung cancer histopathology images using deep learning. Nat Med. 2018;24(10):1559–1567.

6. Campanella G, Hanna MG, Geneslaw L, et al. Clinical-grade computational pathology using weakly supervised deep learning on whole slide images. Nat Med. 2019;25(8):1301–1309.

7. Chen RJ, Ding T, Lu MY, et al. Towards a general-purpose foundation model for computational pathology. Nat Med. 2024;30(3):850–862.

8. Vorontsov E, Bozkurt A, Casson A, et al. A foundation model for clinical-grade computational pathology and rare cancers detection. Nat Med. 2024;30(10):2924–2935.

9. Xu H, Usuyama N, Bagga J, et al. A whole-slide foundation model for digital pathology from real-world data. Nature. 2024;630(8015):181–188.

10. 10. Saillard C, Jenatton R, Llinares-López F, et al. H-optimus-0. Bioptimus; 2024. https://github.com/bioptimus/releases/tree/main/models/h-optimus/v0

11. Ilse M, Tomczak J, Welling M. Attention-based deep multiple instance learning. In: Proceedings of the 35th International Conference on Machine Learning. PMLR 80; 2018:2127-2136.

12. Shao Z, Bian H, Chen Y, et al. TransMIL: transformer based correlated multiple instance learning for whole slide image classification. In: Advances in Neural Information Processing Systems 34; 2021.

13. Zhang J, Nguyen AT, Han X, Trinh VQ, Qin H, Samaras D, Hosseini MS. 2DMamba: efficient state space model for image representation with applications on giga-pixel whole slide image classification. In: Proceedings of the IEEE/CVF Conference on Computer Vision and Pattern Recognition; 2025:3583-3592.

14. Pati P, Jaume G, Foncubierta-Rodríguez A, et al. Hierarchical graph representations in digital pathology. Med Image Anal. 2022;75:102264. doi:10.1016/j.media.2021.102264

15. Hosein AN, Brekken RA, Maitra A. Pancreatic cancer stroma: an update on therapeutic targeting strategies. Nat Rev Gastroenterol Hepatol. 2020;17(8):487–505.

16. Jain S, Wallace BC. Attention is not explanation. In: Proceedings of the 2019 Conference of the North American Chapter of the Association for Computational Linguistics; 2019:3543-3556.

17. Hörst F, Rempe M, Becker H, Heine L, Keyl J, Kleesiek J. CellViT++: energy-efficient and adaptive cell segmentation and classification using foundation models. Comput Methods Programs Biomed. 2026;277:109206.

18. Koh PW, Nguyen T, Tang YS, et al. Concept bottleneck models. In: Proceedings of the 37th International Conference on Machine Learning. PMLR 119; 2020:5338-5348.

19. Moffitt RA, Marayati R, Flate EL, et al. Virtual microdissection identifies distinct tumor- and stroma-specific subtypes of pancreatic ductal adenocarcinoma. Nat Genet. 2015;47(10):1168–1178. doi:10.1038/ng.3398

20. Akbar AR, Levya A, Esnakula A, et al. Inferring clinically relevant molecular subtypes of pancreatic cancer from routine histopathology using deep learning. medRxiv [Preprint]. 2026:2026.01.06.26343539. doi:10.64898/2026.01.06.26343539

21. Lu MY, Williamson DFK, Chen TY, Chen RJ, Barbieri M, Mahmood F. Data-efficient and weakly supervised computational pathology on whole-slide images. Nat Biomed Eng. 2021;5(6):555–570.

22. Schürch CM, Bhate SS, Barlow GL, et al. Coordinated cellular neighborhoods orchestrate antitumoral immunity at the colorectal cancer invasive front. Cell. 2020;182(5):1341–1359.e19. doi:10.1016/j.cell.2020.07.005

23. Wang LM, Silva MA, D’Costa Z, et al. The prognostic role of desmoplastic stroma in pancreatic ductal adenocarcinoma. Oncotarget. 2016;7(4):4183–4194. doi:10.18632/oncotarget.6770

24. Suzuki S, Nagakawa Y, Miyamoto R, et al. Prognostic factors based on histological categorization of desmoplastic reactions in pancreatic cancer. World J Surg. 2024;48(8):1973–1980. doi:10.1002/wjs.12269

